# Safe clinical practice for patients hospitalised in mental health wards during a suicidal crisis: a qualitative study of patient experiences

**DOI:** 10.1101/2020.05.01.20087916

**Authors:** Siv Hilde Berg, Kristine Rørtveit, Fredrik A. Walby, Karina Aase

## Abstract

**Aim:** The aim of this study was to explore suicidal patients’ experiences of safe clinical practice during hospitalisation in mental healthcare. The study was guided by the following research question: How do suicidal patients experience safe clinical practice during hospitalisation in mental health wards?

**Design, setting and participants:** A qualitative design with semi-structured individual interviews was applied. Eighteen patients hospitalised with suicidal behaviour in specialised mental healthcare for adults at a Norwegian hospital participated in the study. Data were analysed thematically and inductively using qualitative content analysis.

**Results:** Patients in a suicidal crisis experienced safe clinical care in mental health wards characterised by the following three themes: (1) being detected by mindful healthcare professionals, (2) receiving tailor-made treatment and (3) being protected by adaptive practice.

**Conclusion:** This study illuminates the experiences of safe clinical practice for patients in a suicidal crisis. The patient group was multifaceted with fluctuating suicidal behaviour, which highlights the importance of embracing personalised activities. Safe clinical practice needs to recognise rather than efface patients’ variability.

**Strengths and limitations of this study:**

• This study used qualitative interviews to provide rich and variable in-depth data of inpatients with suicidal behaviour, which is an under-researched group.
• Patient experience consultants were involved in the design of the study.
• The study results are suitable for conceptual development of safe clinical practice for suicidal patients.
• The patient sample provided rich variability regarding diagnoses, symptom/function level, sex, number of previous hospital admissions and compulsory/voluntary admissions.
• The qualitative methodological approach is not suited for assessing the effects of interventions.

## Background

Patients in mental health wards are a population at particular risk of suicide (1, 2). Inpatient suicide constitutes a proportionately small but clinically important fraction of suicides, and it is a major issue for patient safety in mental inpatient care (3). How to define and understand patient safety in mental inpatient care has been rarely explored (4, 5). Patient safety in mental healthcare is commonly described in physical terms (5). However, other topics emerge when suicidal patients’ experiences are considered. In a systematic review (6), we found that suicidal inpatients felt safe due to their connection with healthcare professionals (HCPs), being protected against their suicidal impulses and through having a sense of control over their lives. No studies have specifically explored what suicidal patients emphasise as vital for their experiences of safety during inpatient care, and the literature on suicidal patients’ experiences of safe clinical practice is limited. Although asking patients at high risk of suicide about suicidal ideations is not associated with increased suicidal ideation (7), knowledge of how suicidal patients experience suicide risk assessments is limited. Suicidal patients’ experiences of being behind locked doors (8) and under constant observation (9, 10) have been sparsely documented in the literature. Although robust evidence supports restricting access to lethal means (11), no studies have explored patients’ experiences of lethal means restriction in hospital wards.

Preventing suicides in wards is a challenging task. Similar to most healthcare activities, safe clinical practice for patients with suicidal behaviour is complex and unpredictable, as knowledge of its underlying principles is incomplete, which often leads to a high degree of uncertainty (12). Expert clinicians cannot predict which patients will commit suicide (13–15), and some patients do not communicate their suicidal ideation to HCPs (8, 16–18). The aetiological heterogeneity of suicidal behaviour further complicates the creation of an all-encompassing model of best treatment practices. Consequently, each patient is understood and approached differently (19). More knowledge on the variability of safe clinical practice from suicidal patients’ perspectives is needed. Thus, this article aims to explore suicidal patients’ experiences of safe clinical practice during hospitalisation in mental healthcare. The study was guided by the following question: How do suicidal patients experience safe clinical practice during hospitalisation in mental health wards?

## Methods

A qualitative design with a phenomenological-hermeneutic approach (20) based on semi-structured individual interviews (21) was applied.

### Setting

The study was conducted at a university hospital in Norway that provides specialised mental health services for patients with mental illness. The hospital treats approximately 10,000 patients per year. Patients were recruited from seven mental health wards for adults: one locked acute ward, one locked specialised ward for affective disorders, four open general mental health wards and one short-term open crisis ward. A national patient safety programme for suicide prevention was taking place at the hospital wards during the data collection. The national programme included a checklist to document whether a patient had been assessed for suicide risk, had received an assessment by a specialist on the first day and had received a safety plan and follow-up appointment at discharge as well as whether the next-of-kin had been contacted (22).

### Participants

The study used a purposeful sampling strategy that aimed to recruit patients with serious suicidal behaviour and/or active suicide ideation who were admitted to open or locked wards in specialised mental health settings for adults. Patients admitted with non-suicidal self-injury were not included in the study (23). The participants were recruited by their therapists at the study sites and self-identified with “being in a suicidal crisis”. The sample consisted of seven men and eleven women (n=18) aged 18-57 years (mean age 40 years). All but one of the participants were of Western origin. See Table 1 for details regarding the participants’ characteristics. A sample size of 18 participants was considered an adequate size to offer sufficient information power to respond to the study aim and ensure participant variability (24).

**Table 1** Participants’ characteristics

### Ethical considerations

All participants provided voluntary and informed consent to participate in the study. They were guaranteed that the information they provided would not be passed on to healthcare professionals in the ward. The interviews were performed before discharge. The timing of the interviews was determined in collaboration with the participants and their therapists to ensure that the participants were sufficiently stable to engage in the interview and without acute suicidal ideation. The study protocol is provided in supplementary file 1 (25). The participants have been given fictitious names here.

### Data collection

The interviews were conducted by the first author (SHB) between September 2016 and January 2017. The interviews were semi-structured and followed an interview guide (supplementary file 2) designed to explore safe clinical practice from different angles. The interview guide was developed in collaboration with an advisory panel and tested in a pilot interview. The pilot interview was included in the study. The interviews focused on the patients’ experiences in the context of daily practices in mental health wards. Of particular interest were interactions with HCPs and experiences of safe clinical practice. A phenomenological-hermeneutic approach was applied during the interviews (20), which implied being sensitive to openness during the interviews by following up with the participant’s responses to the guided questions (20). The interviews lasted for a median of 70 min. The first author (SHB) transcribed the interviews verbatim.

### Analysis

The data were analysed using a phenomenological-hermeneutic approach to content analysis, which guided a systematic move from the manifest content towards a higher level of abstraction and interpretation (26, 27). Each interview transcript was read several times by SHB to gain an overall understanding of what the participant expressed. Collaborative discussions of first impressions were conducted with the authors KR and KAA. The unit of analysis was related to experiences of safe clinical practice across the entire data set. These units were marked and condensed by SHB. In an attempt to understand the life world of each individual, the meaning units pertaining to each participant were condensed and coded separately before moving to more general codes across the data set (20). At this stage of analysis, the manifest content was coded (27). The codes were sorted into five content areas that shed light on specific aspects (talking about suicide, recognising acute suicidality, relational interactions and the therapeutic milieu, protection and treatment). Categories representing a thread through the codes were created using tables and abstracted into three themes and seven sub-themes. The analytical process constantly moved between the whole and the parts (20). The authors read and reread the text to grasp the meaning in relation to the study’s aim and to determine the meaning of the data for the participants. The interpretations and findings were continuously discussed by the authors, and feedback on the themes was provided by the advisory panel, which increased reflexivity and allowed interpretations to be contested and nuanced (28).

## Results

All participants had active suicidal ideation during inpatient care, and nine had recently attempted suicide prior to their admission to mental healthcare. Safe clinical practice for suicidal inpatients was described by three themes with nine sub-themes, as displayed in Table 2.

**Table 2.** Themes and sub-themes

| Themes | Being detected by mindful HCPs | Receiving tailor-made treatment | Being protected by adaptive practice |
| --- | --- | --- | --- |
| Sub-themes | Struggle to communicate suicidal ideations<br>Sensitivity towards deterioration<br>Understood in trusted and familiar relationships | Relieved emotional pressure<br>Collaborative dialogue | Withdrawing from and mastering the outside world<br>Internal and external control<br>Closeness and distance during observation |

### Being detected by mindful HCPs

Patients experienced safe clinical practice when *being detected by mindful HCPs* during acute suicidal deteriorations. As they struggled to communicate their suicidal ideation, they were recognised by HCPs, who showed sensitivity towards their deterioration. Their suicidal behaviour was better understood in trusted and familiar relationships.

#### Struggle to communicate suicidal ideations

Several participants found it difficult to verbalise their suicidal ideation, which they experienced as more profound during episodes of severe mental illness. This experience was related to losing the ability to articulate their inner thoughts when mentally ill, a fear of being locked inside a mental ward, being fixated on death, or having suicidal impulses with sudden deteriorations and acting on impulse without telling anyone. They depended on others to recognise and express their psychological needs when they deteriorated. Family members fulfilled this function before admission, and HCPs did so in the ward: *“I did not say so much (about my suicidal ideation) at the beginning. It was them (parents and girlfriend) who explained most of it because I did not manage to talk. I was completely broken down.”* (Nathan)

Because they were limited by fear, mental illness and difficulty with verbal expression, many of the participants stated that the severity of their suicidal ideation was never detected during formal risk assessments.

Many participants felt unsafe when they were hospitalised through the emergency room and the centralised acute ward because of reduced predictability in terms of whom they would meet and where they would be transferred next. For some of the participants, in particular those admitted for the first time, this insecurity prevented them from verbally communicating their suicidal ideation and reaching out to HCPs for help, as they feared being misunderstood, misinterpreted or mistreated in the form of punishment or seclusion.

#### Sensitivity towards deterioration

Participants experienced that HCPs showed sensitivity towards their acute suicidal state, which saved them from an impending suicide attempt. The HCP who responded was not always the participant’s contact person. The situations were described as “being picked up” or “being read” by someone who was mindful, who cared about them as an individual, who was vigilant and who was able to immediately make sense of changes in their mental state by reading their body language, signs of instability or signs of withdrawal. Patients experienced being seen beyond spoken words by HCPs who acted as lifeguards; they noticed and heard everything: *“There is one nurse who reads me like an open book. She picked me up and managed to read me so clearly and get hold of me. Her presence prevented suicide…She says that she can see it in my face, my eyes and my body posture and that I start tightening my fists.”* (Aina)

The participants experienced that the HCPs immediately understood how to change their suicidal mind-set through, among other strategies, talking about casual everyday topics, addressing sleep problems, connecting and showing genuine interest, thus helping them to regulate their emotions.

Some participants also described that they required HCPs to interpret their spoken words as they struggled to use the term “suicidal” when communicating suicidal ideations, e.g., *“I am in pain; I need to go out for a walk”* (Aina) and *“My life is truly hard to live”* (Ester). In another example, when Patricia said, *“Just send me home; there is nothing here that works for me”*, she planned to go home and take pills to commit suicide, but a nurse understood her communication and told her that she had been neglected in the ward and that she should be taken seriously. Patricia expressed that this understanding saved her from an impending suicide attempt.

#### Understood in trusted and familiar relationships

The participants sought trusted and familiar relationships in the healthcare system because such relationships gave them predictability in terms of how their suicidal behaviour would be understood and treated. Participants who had been hospitalised previously described active strategies for being admitted to a familiar ward milieu. The safety plan helped them to be hospitalised in a familiar place. Being in a familiar place was emphasised as vital for the detection of acute deterioration because such familiarity meant that the participants were close to HCPs who knew from experience how the patient deteriorated and how to intervene: *“They know me, and that is why I think it is important to be admitted to the same ward. They have seen it in the change in my mental state, the things I say and do not say, my facial expressions. They have read me when I get truly, truly silent; then I am ill, and they watch me extra carefully… I have survived because they have watched me like hawks. They have given me my personal freedom, but not too much.”* (Gunn)

The participants described the active strategies that they used to cope with their suicidal deteriorations when they did not have access to HCPs who they perceived as being able to read their suicidal behaviour fluctuations. Turid described how she was saved from suicide attempts by fellow patients who detected her behaviour and called ward personnel at times when she deteriorated and by ensuring that she used medications to fall asleep in order to keep her safe from her impulses at night. These strategies were experienced as unfortunate and made the participants feel unsafe, as they were used to compensate for the lack of trusted HCPs in the ward.

#### Receiving tailor-made treatment

Safe clinical practice was experienced when *receiving tailor-made treatment*, which relieved emotional pressure through targeting underlying stressors and mental health issues. A collaborative dialogue was preferred during suicide risk assessment.

##### Relieved emotional pressure

The participants presented diverse reasons for their suicidal behaviour, which were approached with equally diverse interventions. When treated as an individual their underlying issues and stressors could be addressed, enabling them to re-establish a feeling of internal emotional control that allowed them to cope with their lives without committing suicide, at least in the short term. Experiences of safe clinical practice were highly related to whether the treatment efficiently relieved emotional pressure. The emotional pressure could be due to chaos in their inner worlds, e.g., difficult feelings, delusions, existential issues and sleep deprivation, and/or the outer world, e.g., relational and economic issues and lack of a place to live. For Eva, her emotional pressure was relieved when she was eventually medicated with a mood stabiliser and her delusions telling her to die faded. For Hannah, her emotional pressure was relieved when she received practical support that helped her cope economically with her new life after surviving a suicide attempt: *“I was very miserable in my job. You are in a prison and they have thrown away the key. The key was the assurance that I would never go back to that job. It gave me hope to live and took away my suicidal thoughts… I felt safe when the social worker guided me in the outer world, because I knew how to take hold of my new life.”* (Hannah).

Their underlying issues were targeted by unique combinations of helpful and lifesaving care at the wards that was tailored to the individual (e.g., psychotherapy, medications, rest, isolation, having a strict daily structure, group therapy and activities) by diverse professionals (e.g., social workers, psychologists, nurses and psychiatrists). When these issues were not addressed, the participants experienced being a great risk to themselves after discharge.

Tailor-made treatment was important to ensuring safe clinical practice for patients with complicated mental health issues, as exemplified by Janet. Janet had a history of trauma due to abuse and felt out of control of her suicidal impulses and flashbacks. She managed to find hope and to cope with her flashbacks by talking about her trapped emotions with a psychologist. However, during acute phases she exhibited a severe lack of self-control, and any attempt to restrain her worsened her flashbacks and suicidality. She managed to gradually improve through treatment with sedatives during acute phases and the presence of HCPs who stayed with her in the bathroom in the dark, as this made her feel safe because no one could find her.

Feeling that the conversation relieved emotional pressure was important when talking about suicide. The participants longed for confirmation that their suffering and suicidal ideation were understandable. Many participants experienced HCPs asking about suicidal ideation, but their pain was not alleviated when they opened up.

> *“They do not have the time, they are looking at their watch, as if they would rather be somewhere else. When they do not take my suicidal ideation seriously, I think I am worthless and should instead keep these thoughts to myself”* (Aina).

Opening up about difficult emotions and suicidal ideation involved being in a vulnerable position, as described by Gunn: *“elaborating on my suicidal thoughts is extremely personal for me. It is worse than undressing and being naked. It is like going to the gynaecologist”*. A lack of emotional confirmation elicited feelings of hopelessness, shame and withdrawal from disclosing suicidal thoughts.

##### Collaborative dialogue

The participants had positive experiences of being assessed for suicide risk when the questions appeared to occur naturally as part of a collaborative dialogue in which they were perceived as individuals and HCPs validated their feelings. Merely asking questions about suicidal ideations was described as “ticking off boxes”, “being a part of a machine”, and “being interrogated”, leading to the impression that their personal experiences, stories and feelings were not important: *“They should ask other questions than just about suicidality, such as what is your life situation like… It is meaningless to be asked about suicidal thoughts and plans when they do not understand the context of why I do not want to live.”* (Kate)

The participants said that when addressing suicidal ideation, the HCPs should tailor their responses and adjust the conversation about suicide towards topics that matter instead of giving only general advice. One example of what was perceived as generic advice was reminding patients to think of their children. However, having children was not necessarily a protective factor for keeping the patients alive at different stages of their suicidal crisis. The participants said that they had periods when they struggled with guilt and felt like a burden and thought that their children would manage better without them. Whether the participants experienced a need to elaborate on their suicidal ideation also varied. While some experienced less suicidal ideation when they shared their inner suicidal thoughts and feelings, others improved by focusing on different topics (e.g., finding hope through coping with economic issues and coping with delusions).

#### Being protected by adaptive practice

Safe clinical practice was experienced when *being protected by adaptive practice* as their suicidal behaviour fluctuated, and the need for protection varied between the participants. Safe clinical practice was experienced as a balance between withdrawing from and mastering the outside world, internal and external control and closeness and distance during observation.

##### Withdrawing from and mastering the outside world

The participants experienced being protected from suicidal impulses during inpatient care by being removed from the overwhelming stressors and demands of the outside world that triggered their suicidal ideation. However, withdrawal was described as a short-term strategy, and they clearly stated they needed to cope with the outside world: “*I struggle with guilt about not coping with things at home. When I am hospitalised, I do not get these reminders all the time and I have fewer episodes of suicidal ideation. At home, I have so much to cope with that the suicidal thoughts are triggered. However, the experience is two-sided: I feel guilty about the fact that I am not with my family and I feel defeated when I do not deal with my home situation because my life should not be here.”* (Ida)

The participants felt safe during discharge when HCPs balanced their need to withdraw from and master the outside world. They needed to feel able to cope with both their symptoms and their life situations to be ready to leave the ward. Safety was also experienced when the participants were involved in the discharge process of finding the right balance between activity and peace, testing this balance during ward leaves and receiving support when the balance failed. The patients emphasised the need for predictability regarding follow-up after discharge for their own safety. They experienced severe anxiety about being discharged without feeling prepared: *“To be notified about discharge on the same day is like hitting the pavement at 100 km per hour. I was discharged without being prepared, and I became very confused and even more of a danger to myself.”* (Gunn)

##### Internal and external control

The participants described experiencing safety from their suicidal impulses through either internal or external control, which changed during their suicidal crisis, as described by Magnus: *“To feel safe from myself, I needed to get out of that psychosis where I believed that I was completely bound to kill myself because I had let everything and everyone down. Because I did not truly want to kill myself…. I lost my sense of self, my motor control, my sight and my concentration during the psychosis. I thought this was the way my life had become… I needed rest, isolation and medication, and with time I understood that I would get better and then I needed to experience that I could function normally again and trust that I wold not kill myself”* (Magnus).

When experiencing safety through external control, the participants felt safe by being physically held back from suicidal impulses, delusions or hallucinations commanding them to commit suicide or moments of overwhelming agitation or despair. Locked doors or restraints replaced their sense of no control, and the lack of such protection placed greater demands on their own self-control. In the aftermath, they perceived that they were being saved from death when they received proper protection: *“Being restrained has a calming effect on me. I can hand control over to others and relax because I know that I cannot do any harm. My suicidal thoughts fade because I know that I am totally without control… When you are so intensely agitated, nothing stops you… Being hospitalised by force has been crucial for not committing suicide.”* (Klaus)

When feeling safe through experiencing internal control, participants had the freedom to experience that nothing happened as a result of their ideations. Barred windows, locked doors and having to walk through metal detectors increased their anxiety regarding losing this freedom and provoked thoughts such as being a *prisoner*, a *child*, or *“having passed the point of no return”*, which was especially evident among those who were admitted for the first time. In such cases, locked wards could result in feelings of claustrophobia, panic attacks and increased suicidal ideation. Patients’ anxiety was reduced when they understood that the physical barriers and procedures were intended to help them.

Being deprived of personal belongings was experienced as a necessary protection for all participants during an acute suicidal crisis, and the procedure was easily accepted and intuitively understood as necessary for their own safety. The participants emphasised the importance of not having access to any potentially lethal items, such as belts or medications, in both open and closed wards to prevent suicide during moments of deterioration.

##### Closeness and distance during observation

Due to the invasiveness of observations, the participants emphasised the need to balance closeness and physical distance. They needed a balance between being acknowledged and seen and being left in peace, having their privacy respected without being given too much freedom: *“Firm but soft, but not too much freedom,”* The participants’ ability to establish relational contact during constant observation varied. Their needs and their ability to connect altered as their mental state fluctuated. Some participants needed active support and dialogue with the HCPs, while others wanted to be left in peace but needed confirmation that the HCPs were present (i.e., outside the room with the door open) if required. Participants experiencing a psychotic episode reported being in a mental state that left them unable to communicate and establish relationships with the HCPs. In this state, they indicated that they simply needed the HCPs to show that they genuinely cared for them, keeping them within sight and recognising their fluctuations. They described being fixated on death and constantly thinking about suicide and therefore experienced the constant presence of HCPs as lifesaving.

Although constant observation was experienced as invasive, in the aftermath of their crisis the participants perceived this practice as safe and necessary to preventing suicide: *“I still hate being followed everywhere when I have a suicide plan, but they watch all the time because they care; it is a sign of humanity. They have saved me many times.”* (Janet).

However, observation was experienced as unsafe when the patients’ need for connection and acknowledgement was neglected and they felt left on their own and ignored. It was important that the HCPs established relationships with the patients and asked how they were doing rather than just *“checking whether they were alive”* and acting as though they were *“guardians of a prison”*. Such practices increased the participants’ suicidality, and for some, this had devastating effects on trust.

When under intermittent observation, patients felt safe by having relationships with HCPs based on trust rather than control. Trusting relationships were established when the participants felt they were treated as valuable and equal human beings. Such encounters could be in the form of simple informal contact, which made the participants feel that the HCPs were available and genuinely cared about them as individuals and were not just doing their job. It made them feel safe knowing that the HCPs would intervene during a suicidal crisis if they were unable to call for help themselves.

## Discussion

This article aimed to explore the experiences of safe clinical practice among patients hospitalised during a suicidal crisis. There was rich variation in the participants’ experiences of safe clinical practice expressed in the following themes: “being detected by mindful HCPs”, “receiving tailor-made treatment” and “being protected by adaptive practice”.

*“Being detected by mindful HCPs”* highlights the experiences of struggling to verbally communicate suicidal ideation, which was more profound during severe mental illness. The connection between the severity of mental illness and the lack of verbal communication of suicidal ideation has been described among patients with depressive disorder (17). Levi-Bels et al. (29) found that suicide attempters who did not verbally communicate suicidal ideation were characterised by higher levels of suicide ideation, distress and victimization than those who did communicate their ideations. An inability to identify and communicate suicidal ideation has also been documented in a sample of patients with psychotic depression (8). Furthermore, the findings of the present study are in line with other findings in the literature that shame and trust issues inhibit honest communication during suicide risk assessment (18, 30) Nevertheless, knowledge regarding how patients who do not communicate their suicidal ideation are saved by others is limited. In the present study, HCPs’ observation of patient behaviour enabled detection of suicidal behaviour in the participants. The study emphasises the importance of understanding warning signs among inpatients (31), in particular for those who struggle to participate in a collaborative dialogue about suicidal ideations. As warning signs vary among the participants in the present study and across time, the success of such understanding seems to be dependent on HCPs who are familiar with and vigilant about changes in a patient’s mental status, irrespective of whether they were that participant’s contact person in the ward. These findings emphasise the importance of a high level of expertise among all HCPs who interact with patients, enabling them to connect with each patient and make sense of her/his situation.

The findings also highlight the importance of being informed about a clear pathway on admission to hospital. The importance of suicidal patients having trust in their HCPs (32–36) has been well documented in the literature. Familiar and trusted relationships are important for enabling suicidal patients to feel safe because they provide predictability in how their suicidal behaviour is understood and approached. Considering that the suicide risk is highest in the first week after psychiatric hospitalisation (37), immediate admission to familiar places that patients trust may be one strategy to employ during re-admissions, as highlighted in the current study.

*“Receiving tailor-made treatment”* highlights the rich variation in underlying issues and associated treatment paths for patients with suicidal behaviour, emphasising that practice is characterised by differing treatment strategies across participants as opposed to practices with high similarity (38), emphasising that suicidal behaviour is characterised by aetiologic heterogeneity (19). The findings indicate that tailor-made treatment efficiently relieved the patients’ emotional pressure through addressing the individuals’ need to re-establish a feeling of control regarding their suicidal impulses. Individualised care and tailored services are highly central topics of patient experiences in healthcare (39); however, their relevance to suicidal patients’ experiences of safety has been less explored. The findings support the assumption that a sense of safety for the individual patient can be achieved through addressing her/his manifestations of suffering, as discussed by Undrill (40). Furthermore, for the suicidal patient, experiences of safety relate to re-establishing a feeling of control, as found by Berg et al. (6).

This study also addresses the how patients experiencing suicide risk assessments as safe. Through the collaborative dialogue and relieving emotional pressure during suicide risk assessment, harm may be avoided, and HCPs may help patients to re-establish a feeling of control. The emphasis on the role of a collaborative assessment of suicide risk that accounts for the suicidal patients’ individual drivers has been described elsewhere (41). Patients have stressed the importance of trust and support to verbally communicate their suicidal thoughts (30, 42). Consequently, this study supports the recommendations provided by the British NICE guidelines (43) to avoid using tools and scales to predict suicide; to manage risk and not merely assess it; and to identify and agree with patients regarding their specific risks (43). Experiencing safety during suicide risk assessments involves a collaborative dialogue, establishing a therapeutic alliance that includes trust, confirmation of feelings and tuning into the patient’s issues to manage emotional pressure. Nevertheless, some patients have difficulties participating in a collaborative dialogue.

The theme *“being protected by adapted practice’’* adds knowledge regarding the dynamic, fluctuating and interactive nature of experiencing protection as a means of safe clinical practice. Patients have utterly different experiences of safety in relation to locked doors, barred windows, restraints and involuntary commitment. This finding is in accordance with other descriptions in the literature, e.g., locked doors have been experienced as both “being admitted to prison” and “having access to shelter” (44), while involuntary commitment has been experienced as both “necessary” and “being cared for” and as “unjust” or a “restriction of autonomy” (45, 46). However, this does not imply that protective interventions are entirely good or bad; it depends on what works for whom (47). It is not a matter of whether doors should be locked but rather which patients need to be behind locked or open doors along with when and how. Locking all wards as a means of safety may have consequences for help-seeking behaviour, compliance and recovery for patients experiencing being safe with internal control. To ensure that healthcare can adjust to the patients’ need for control, it is necessary to have both open and locked wards. Furthermore, identifying patients who suffer emotionally when they are physically protected is important to minimising their catastrophic thoughts and emotional reactions. To our knowledge, this is the first study of patients’ experiences of being deprived of lethal means in hospital wards. There is robust evidence for the preventive effect of not having access to any lethal means in hospital wards (11), and this study provides evidence that patients do not perceive this procedure as invasive when they understand its purpose.

Safe clinical practice was additionally a matter of having a balance between closeness and distance during observation. The importance of supportive HCPs who acknowledge patients during constant observation (10, 48, 49) and interchange between control and building the therapeutic relationship (50) has been described in previous research. This study adds to the importance of understanding the dynamic relationship during observation. Patients’ needs change throughout a suicidal crisis, so to their capability to connect with others. Safe clinical practice involves a flexible relationship during observations, where HCPs tune into patients’ need for closeness and distance. During this complex endeavour, HCPs can make a difference between life and death. Both experiencing inattentive HCPs and feeling ignored can potentially increase suicidal behaviour and cause patients to feel unsafe. Accordingly, this study supports the perspective of Cutcliffe and Barker (51) that observation should be regarded as a dynamic relational practice, without neglecting the vitality of being watchful and physical present.

### Strengths and weaknesses of the study

A phenomenological-hermeneutic approach was employed with a sample of 18 participants. While the methodological approach cannot study the effects of interventions, it can provide a deeper understanding of how safe clinical practice is experienced and how it varies among patients. Credibility is strengthened by including a sample that covered significant variations, and participants with relevant experiences with the phenomenon under study (26), and through providing a sample size with sufficient information power (24). The findings cannot be generalised to the entire population of patients hospitalised in mental health wards during a suicidal crisis. Nevertheless, theoretical generalisations can be made regarding the suicidal patients’ perspectives on safe clinical practice.

## Conclusion

This study contributes to the understanding of how suicidal patients experience safe clinical practice. Safe clinical practice is experienced by patients hospitalised during a suicidal crisis when they are detected by mindful HCPs, receive tailor-made treatment and are protected by adaptive practice. The patient group was multifaceted with fluctuating suicidal behaviour, which highlights the importance of embracing personalised activities. Safe clinical practice needs to recognise rather than efface patients’ variability. This requires expert knowledge from HCPs in terms of interpersonal skills, competence and experience with understanding mental illness and how to adapt practices to the individual patient.

## Data Availability

The datasets generated and/or analysed during the present study are not publicly available due to restrictions regarding individual privacy, but anonymised data are available from the corresponding author upon reasonable request.

## Acknowledgements

We would like to thank the advisory panel members for this study who contributed feedback on the recruitment strategies, the interview guides and the manuscript draft: Dag Lieungh (patient experience consultant), Målfrid J. Frahm Jensen (patient experience consultant), Gudrun Austad (inpatient and community suicide prevention; mental health nurse), Kristin Jørstad Fredriksen (consultant psychiatrist), Camilla Hanneli Batalden (consultant clinical psychologist), Liv Sand (consultant clinical psychologist) and Sigve Dagsland (consultant clinical psychologist).

## Supplementary files

1. Study protocol
2. Interview guide

## Patient consent for publication

Not required.

## Ethical approval

This study was approved by the Regional Committees for Medical and Health Research Ethics (2016/34; Norway).

## Author contributions

SHB conceived and designed the study and collected and organised the data. SHB, KR and KAA read transcripts and participated in analytical reflections and validation of the analysis. SHB drafted the manuscript, and all authors provided critical revisions for intellectual content.

## Author information

SHB: PhD scholar in Health and Medicine, consultant clinical psychologist, safety science. KR: PhD, researcher in nursing science, mental health nurse. FAW: Consultant clinical psychologist, researcher in suicide research and head of the Norwegian Surveillance System for Suicide in Mental Health and Substance Misuse Services. KA: professor in patient safety, researcher in health services research.

